# Associations between mosaic loss of sex chromosomes and incident hospitalization for atrial fibrillation in the United Kingdom

**DOI:** 10.1101/2024.05.29.24308171

**Authors:** Jungeun Lim, Aubrey K. Hubbard, Batel Blechter, Jianxin Shi, Weiyin Zhou, Erikka Loftfield, Mitchell J. Machiela, Jason Y.Y. Wong

## Abstract

**Background:** Mosaic loss of chromosome Y (mLOY) in leukocytes of men reflects genomic instability from aging, smoking, and environmental exposures. A similar mosaic loss of chromosome X (mLOX) occurs among women. However, the associations between mLOY, mLOX, and risk of incident heart diseases are unclear.

**Methods:** We estimated associations between mLOY, mLOX, and risk of incident heart diseases requiring hospitalization, including atrial fibrillation, myocardial infarction, ischemic heart disease, cardiomyopathy, and heart failure. We analyzed 190,613 men and 224,853 women with genotyping data from the UK Biobank. Among these participants, we analyzed 37,037 men with mLOY and 13,978 women with mLOX detected using Mosaic Chromosomal Alterations caller. Multivariable Cox regression was used to estimate hazard ratios (HRs) and 95% confidence intervals (CIs) of each incident heart disease in relation to mLOY in men and mLOX in women. Additionally, Mendelian randomization (MR) was conducted to estimate causal associations.

**Results:** Among men, detectable mLOY was associated with elevated risk of atrial fibrillation (HR=1.06, 95%CI:1.03-1.11). The associations were apparent in both never-smokers (HR=1.07, 95%:1.01-1.14) and ever-smokers (HR=1.05, 95%CI:1.01-1.11) as well as men > and ≤60 years of age. MR analyses supported causal associations between mLOY and atrial fibrillation (HR_MR-PRESSO_=1.15, 95%CI:1.13-1.18). Among post-menopausal women, we found a suggestive inverse association between detectable mLOX and atrial fibrillation risk (HR=0.90, 95%CI:0.83-0.98). However, associations with mLOY and mLOX were not found for other heart diseases.

**Conclusions:** Our findings suggest that mLOY and mLOX reflect sex-specific biological processes or exposure profiles related to incident atrial fibrillation requiring hospitalization.

**WHAT IS KNOWN?:**

- A previous population study found links between death from atrial fibrillation and heart failure, and mosaic loss of chromosome Y (mLOY) in leukocytes, which is a marker of genomic instability, environmental exposures, and aging. Additionally, mLOY has been associated cross-sectionally with prevalent cardiovascular and metabolic diseases.
- A similar but less common mosaic loss of chromosome X (mLOX) occurs among women but is role in disease pathogenesis is less characterized.
- The contributions of mLOY and mLOX to risk of hospitalization for incident atrial fibrillation, heart failure syndrome, and other heart diseases are unclear.

**WHAT THE STUDY ADDS?:**

- Among men, mLOY was associated with elevated risk of incident atrial fibrillation. Further, we found suggestive evidence linking mLOX to atrial fibrillation risk among women. Taken together, mLOY and mLOX potentially reflects sex-specific factors related to the pathogenesis of atrial fibrillation.

## Introduction

The Y chromosome contains genes that play crucial roles in biological pathways involved in male sex determination, spermatogenesis, and sexual maturation. Further, genetic variants on the Y chromosome have been linked to fundamental cellular housekeeping processes including mitosis, cell cycle regulation, DNA damage detection and repair, and apoptosis ^1^. As such, alterations to the dosage of genes located on the Y chromosome can potentially influence the pathogenesis of chronic diseases that involve these underlying biological pathways. Mosaic loss of chromosome Y (mLOY) is a common genomic alteration that manifests in a fraction of cells as men age. Depending on the study and calling methods, an estimated 7-57% of older men, generally over 65 years of age, have detectable mLOY in circulating leukocytes ^1–7^. Variation in mLOY has both genetic and environmental components, having associations reported with cigarette smoking, outdoor air pollution, and arsenic ^1, 4, 7, 8^.

In population studies, mLOY has been linked to risk of various chronic diseases ^9–12^ ^13^ ^14^; however, investigations into the contribution of mLOY to the pathogenesis of cardiovascular diseases (CVDs) are still in early stages ^14, 15^. In the UK Biobank, mLOY in leukocytes was found to be cross-sectionally associated with prevalent diabetes and heart disease ^15^.

Furthermore, recent analyses found that mLOY was associated with increased risk of death from both atrial fibrillation and heart failure syndrome ^16^. The biological mechanisms underlying these relationships with heart diseases in humans remain unclear. However, studies have suggested links between mLOY and increased total and bioavailable testosterone^17^, metabolic biomarkers that we recently found to be associated with incident hospitalization for heart failure^18^.

The X chromosome contains essential housekeeping genes responsible for basic biological function ^19^. Among men, mosaic loss of chromosome X (mLOX) in leukocytes are poorly tolerated and rarely observed ^20^. Among women however, mLOX in leukocytes can change the dosage and allelic balance of genes on the maternal or paternal X chromosome. Given that one X chromosome is epigenetically inactivated in female diploid cells, the biological impact of mLOX is determined by which X-chromosome is lost ^21^. Previous analyses found that mLOX preferentially affects the inactivated X chromosome ^19^. Recent meta-analyses of large genome-wide association studies (GWAS) conducted in multiple biobanks have identified 49 independent genetic variants associated with mLOX ^21^. However, little is known about its influence on the risk of heart diseases.

Despite a growing body of literature on the relationships between mosaic loss of sex chromosomes and prevalent CVD and CVD-related mortality, their associations with disease incidence are unclear. Investigating the impact of mosaic loss of sex chromosomes in leukocytes on risk of incident heart diseases is important for understanding the role of genomic instability and immune function in disease pathogenesis and could potentially have translational relevance towards risk prediction. To address these gaps in knowledge, we leveraged the extensive demographic, clinical, and GWAS data from the UK Biobank. In a population free of CVD at baseline, we evaluated the prospective associations between pre-diagnostic leukocyte mLOY among men, mLOX among women, and risk of incident atrial fibrillation, myocardial infarction, ischemic heart disease, cardiomyopathy, and heart failure syndrome requiring hospitalization.

## Methods

### Study design

The UK Biobank is a prospective cohort study and has been described in detail (http://www.ukbiobank.ac.uk/) ^22, 23^. Briefly, the source population included adults aged 40-69 years who lived ≤40 km of 22 study assessment centers across England, Wales, and Scotland. Nearly 9.2 million people registered in the National Health Service (NHS) were mailed invitations to participate in the study, while 503,317 (5.5%) visited the assessment centers in 2006-2010 and were enrolled ^22^. The volunteers completed touchscreen questionnaires and physical examinations, and provided blood samples for bioassays and genomic analyses. The UK Biobank is regularly updated and our dataset from August 2022 included 502,409 subjects (project number: 28072).

### Genome-wide association study (GWAS) data

GWAS data was available for 488,377 UK Biobank participants as described ^24^. Briefly, genotyping was performed by the Affymetrix Research Services Laboratory (now part of Thermo Fisher Scientific) on two similar custom arrays. Among these subjects, 49,950 were scanned on the UK BiLEVE Axiom array, while the remaining 438,427 participants were scanned using the UK Biobank Axiom array. The GWAS dataset combines results from both arrays and there were 805,426 markers in the released genotype dataset. Approximately ∼3% of eligible participants who consented to genetic analyses did not have sufficient extracted DNA to conduct the genotyping. Affymetrix used a custom genotype calling pipeline and the genotype data were quality controlled (QC). Furthermore, the dataset was phased and nearly 96 million genotypes were imputed using computationally efficient methods combined with the Haplotype Reference Consortium and UK10K haplotype resources. Information on the QC pipeline and population structure and relatedness have been described in detail ^24^.

### Detection of mosaic loss of sex chromosomes

Raw genotyping array intensity data were used to calculate B allele frequencies (BAFs) and log_2_ R ratios (LRR), which were then analyzed using MoChA v2022-01-12 (https://github.com/freeseek/mochawdl) to detect mosaic chromosomal alteration (mCA) regions as previously described ^25–27^. The mCA calling was performed by Giulio Genovese (Broad Institute of MIT and Harvard). MoChA uses Viterbi hidden Markov models to integrate LRR and BAFs, leveraging haplotype information to detect subtle imbalances between maternal and paternal allelic fractions in a cell population. SHAPEIT4 software was used for phasing to infer haplotypes. LRR was used to determine the status of events (i.e., gain, loss, copy neutral loss of heterozygosity). We focused on subjects with a sample call rate >=0.97 and baf_auto <=0.03 to ensure high-quality samples for subsequent analysis.

Post-processing of the mCA data to generate the mLOX and mLOY calls was performed at the National Cancer Institute. For mLOY calling, we identified male subjects exhibiting mLOY based on the established criteria outlined at (https://github.com/freeseek/mocha). These criteria included a minimum event size > 2MB and a relative copy number (rel_cov) < 2.5 in the pseudoautosomal region 1 (PAR1) region. The proportion of cells with Y-chromosome loss (“mosaic cell fraction”, CF) was estimated using BAF values in the mLOY region. To improve mLOY detection, mosaic fraction estimates for Y-chromosome events were derived using BAF deviation on the pseudoautosomal 1 region. For the mLOX calling, we identified female subjects exhibiting mLOX based on the standard criteria specified at (https://github.com/freeseek/mocha), including a minimum event size >100MB and a relative copy number (rel_cov) < 2.5 on the X chromosome. The CF with X-chromosome loss was estimated using BAF values in the mLOX region. Mosaic CF has emerged as an important consideration in mCA analyses, as increased proportions reflect greater white blood cell aberrations and stronger disease associations have been found at CFs >10% ^28^.

### Ascertainment of heart disease cases

Heart disease cases were defined using in-patient hospital diagnoses coded according to International Classification of Disease (ICD) versions 9 and 10. Atrial fibrillation was defined using ICD-10 (I48, I48.0, I48.1, I48.2, I48.3, I48.4, I48.9) and ICD-9 (427.3). Myocardial infarction was defined using ICD-10 (I21, I22, I23) and ICD-9 (412). Ischemic heart disease / angina pectoris was defined using ICD-10 (I20, I24, I25) and ICD-9 (411.1, 413.0, 413.1, 414.0, 414.8, 414.9). Cardiomyopathy was defined using ICD-10 (I25.5, I42.0, I42.5, I42.9, I43) and ICD-9 (425.4). Heart failure syndrome was defined using ICD-10 (I42.8, I42.1, I50, I50.1, I50.9, I11.0, I13.0, I13.2) and ICD-9 (428.0, 428.1).

We conducted sensitivity analysis using a smaller subgroup of UK Biobank participants (n∼ 229,972, 45% of study population) in which preliminary raw primary care data were collected (https://biobank.ndph.ox.ac.uk/showcase/showcase/docs/primary_care_data.pdf). These primary care data presumably capture more moderate cases that did not initially require hospitalization. Importantly, there is no national system for collecting or sharing primary care data and the UK Biobank has worked with various data suppliers and other intermediaries to obtain primary care data. Participants provided written consent for linkage of primary care data to their health-related records. Here for example, we defined atrial fibrillation using Clinical Terms Version 3 codes that correspond to a text search for “atrial fibrillation”. In the subset with primary care data, we analyzed incident outcomes as: 1) general practitioner diagnosed specific heart disease, 2) hospitalization for specific heart disease, and 3) combined general practitioner and hospitalization for specific heart disease.

### Inclusion / exclusion criteria

Among the 502,409 participants at baseline, we excluded 372 participants with discrepancy between genetic and self-reported sex, 40 who dropped out of the study, 2257 with prevalent heart failure syndrome and 23,199 with prevalent CVD, which left 476,541 participants. Among these subjects, 463,082 had genotype data for detecting mLOY or mLOX (Table 1) and were considered for further analysis.

**Table 1:**
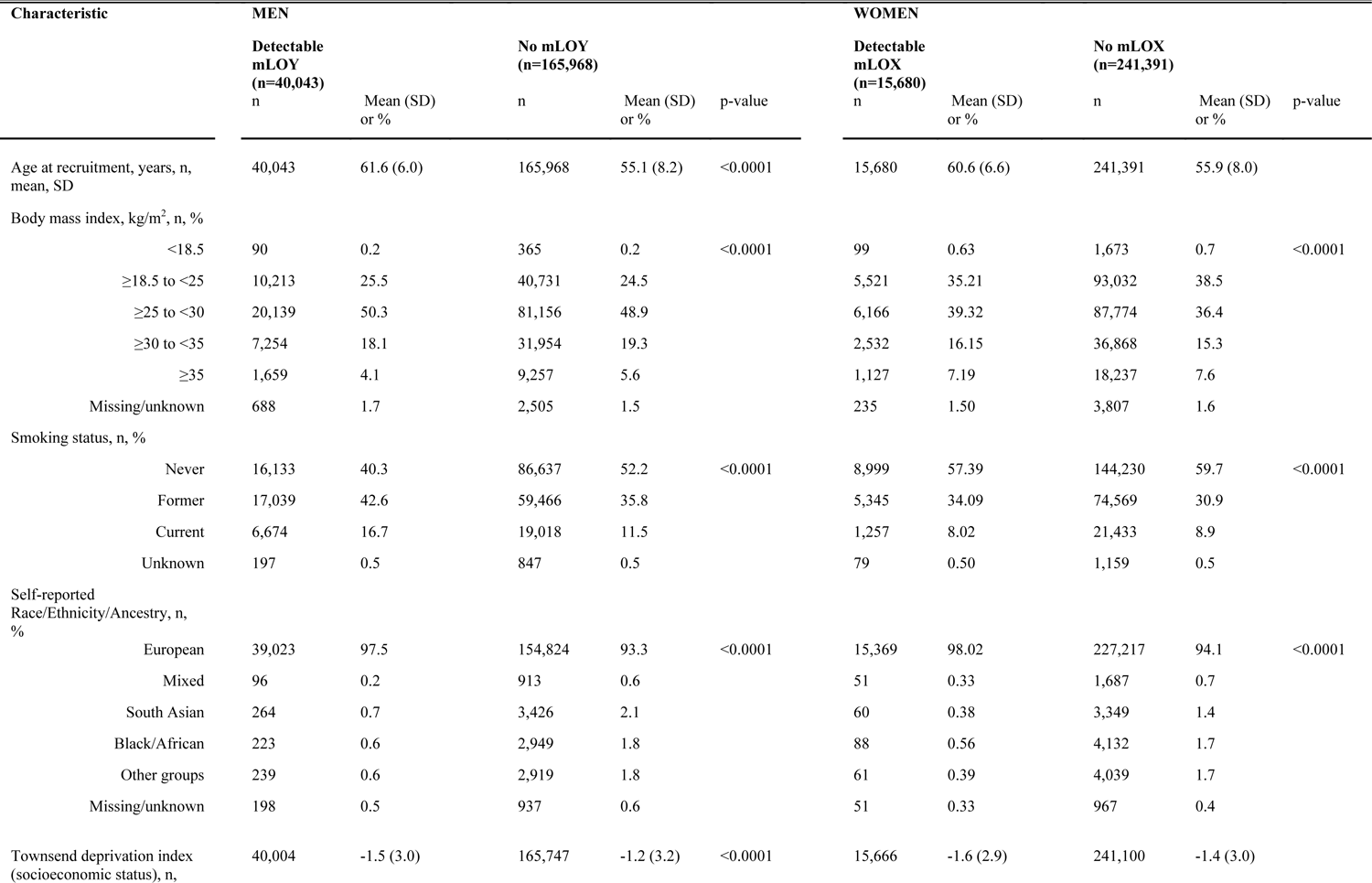
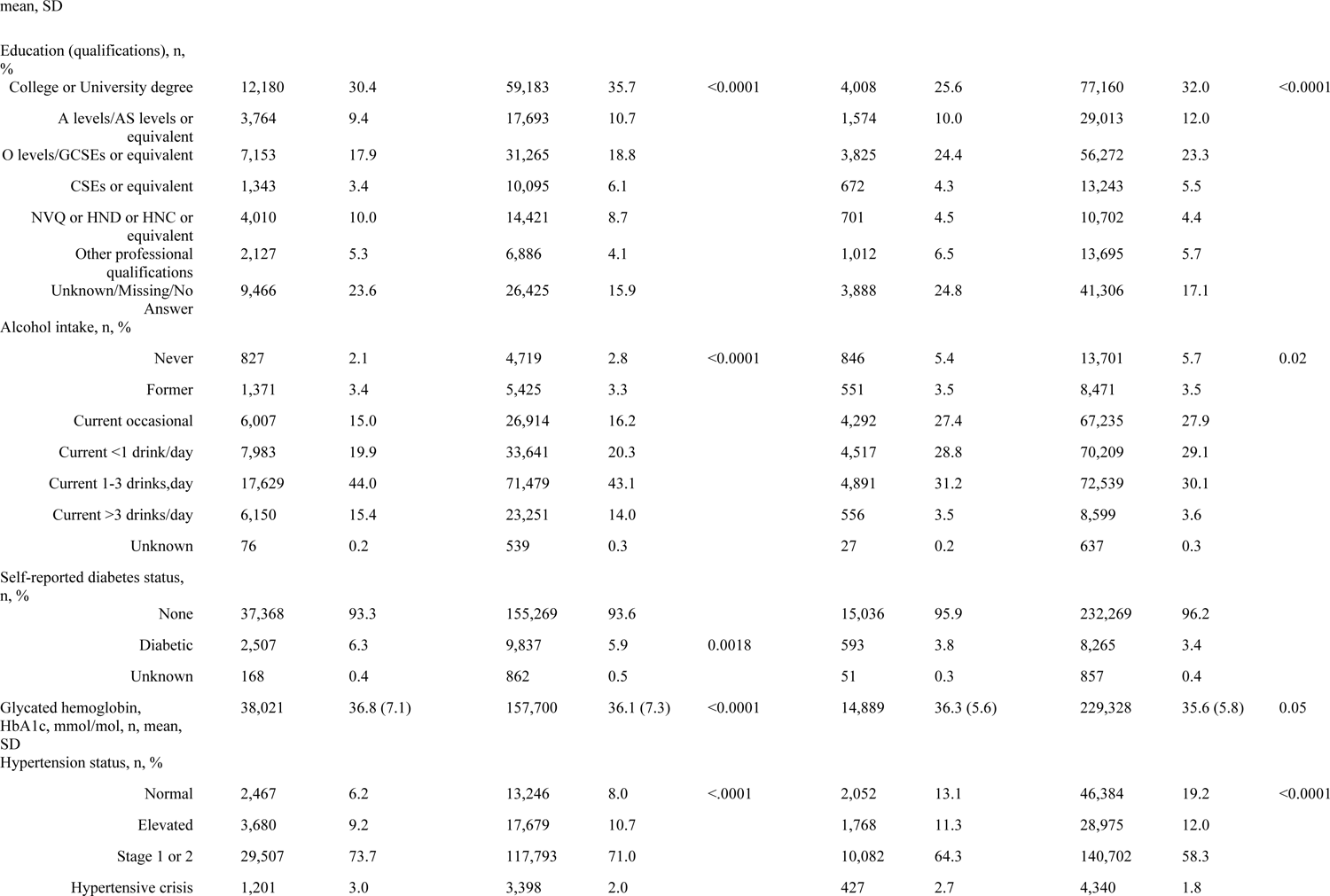
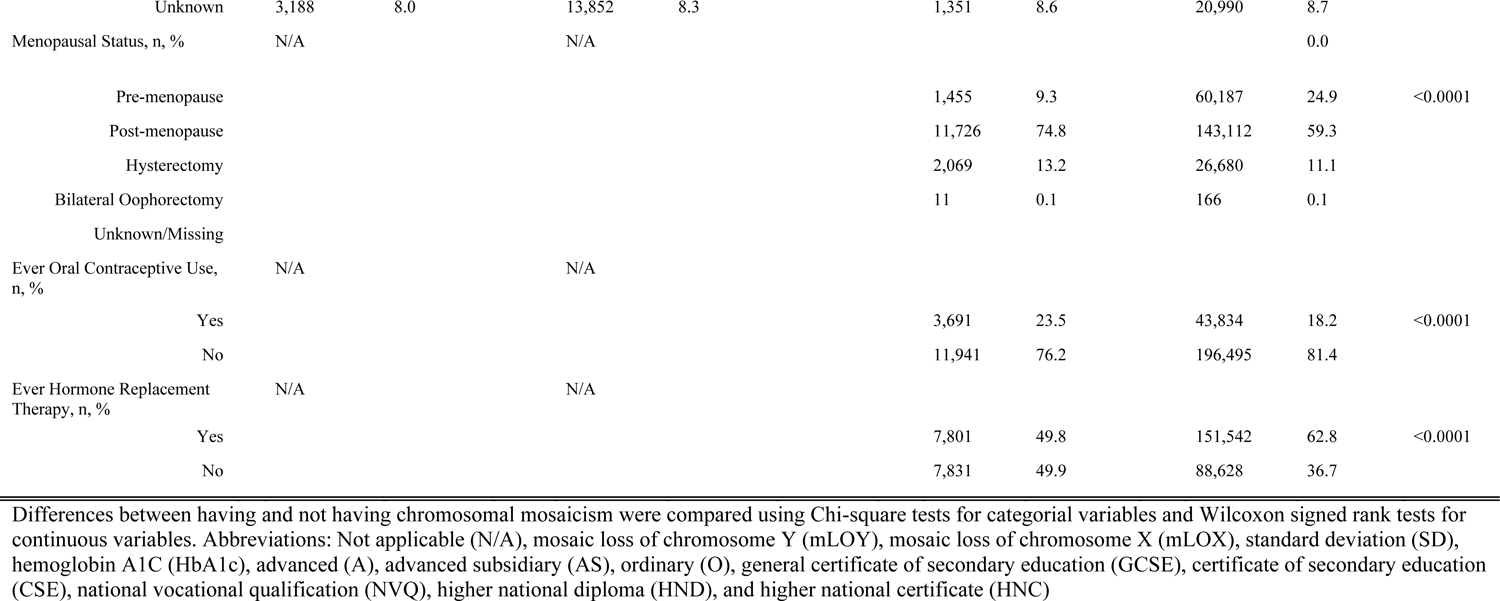
Baseline characteristics of UK Biobank study population with and without mosaic loss of sex chromosomes (n=463,082)

### Follow-up

For each participant, the prospective follow-up started at the date of visit to the assessment center in 2006-2010 and ended at the date of first incident diagnosis (hospitalization) of heart disease, death, or administrative censoring (i.e., September 20^th^, 2021, for England and Wales and October 31^st^, 2021, for Scotland), whichever came first. Vital status and the primary underlying cause of death for participants were provided by the NHS Information Centre and the NHS Central Register. The UK Biobank study was approved by the National Information Governance Board for Health and Social Care and the NHS North West Multicenter Research Ethics Committee. All participants provided electronic informed consent.

### Main analyses

Separate multivariable Cox regression models were used to estimate hazard ratios (HR) and 95% confidence intervals (CI) of incident hospitalization for atrial fibrillation, myocardial infarction, ischemic heart disease / angina pectoris, cardiomyopathy, and heart failure syndrome in relation to mLOY (detectable vs. not detectable) among men and mLOX (detectable vs. not detectable) among women. For mLOY analyses among men, we adjusted for potential confounders including study assessment center, age at recruitment (continuous), self-identified race/ethnicity to indirectly adjust for culture-specific dietary patterns, environmental factors, social determinants of health, etc. (European, Black/African, mixed, South Asian, and other groups), 10 principal components for genetic ancestry, educational attainment (i.e., qualifications), smoking status (never, former, current), body mass index (BMI; <18.5, ≥18.5 to <25, ≥25 to <30, ≥30 to <35, and ≥35 kg/m^2^), material deprivation (Townsend deprivation index, socioeconomic status, continuous), alcohol intake (never, former, current occasional, current <1 drink/day, current 1-3 drinks/day, current >3 drinks/day, unknown), self-reported diabetes status (none, diabetic, unknown), glycated hemoglobin (HbA1c, mmol/mol, continuous), hypertension status based on average systolic and diastolic blood pressure at baseline (normal, elevated, stage 1 and 2 hypertension, hypertensive crisis, and unknown), total white blood cell count at enrollment, and any cancer diagnosis at enrollment. For mLOX analyses among women, we adjusted for the same covariates as men in addition to oral contraceptive use (never and ever), hormone replacement therapy use (never and ever), and menopausal status (pre-menopause, natural post-menopause, artificial menopause, and missing/unknown). We conducted additional analyses stratified by leukocyte mosaic CFs, smoking status (never and ever), age groups (> and ≤60 years), menopausal status (pre- and post-menopause) among women, as well as a sensitivity analysis restricted to European genetic ancestry participants. We analyzed participants with complete data on independent variables. Overall and within stratified and subgroup analyses, p-values <0.05 were considered noteworthy and those below a Bonferroni-corrected α-threshold of 0.01 were considered statistically significant (i.e., 0.05 divided by a family of 5 tests corresponding to each heart disease outcome).

### Attenuation and Sensitivity Analyses

To tightly control potential residual confounding from smoking in the main analyses, we adjusted for a 27-category variable for smoking history and intensity ^12, 15^. Additionally, to account for non-linear relationships with age, we included polynomial terms for age in the models.

### Mendelian Randomization analyses

Among the measured mLOY, mLOX, and heart disease associations found to be statistically significant in the Cox regression analyses, we further assessed potential causal associations using several established Mendelian randomization (MR) methods. First, we used the inverse variance method (IVW), an effective approach when all included genetic variants are valid instrumental variables (IVs) ^29^. However, when horizonal pleiotropy is present, the estimate from IVW approach can be potentially biased with over-estimation ^29, 30^, and we therefore employed additional approaches as sensitivity analyses. We used the weighted median approach, which has been shown to produce valid causal estimates if the invalid IVs account for <50% of the weight of the studies ^29^. Next, we used MR-Egger, which unlike other MR methods, estimates the dose-response relationship between the genetic associations with mLOY and those with the heart disease outcome ^31^. A non-zero intercept from MR-Egger regression indicates potential horizontal pleiotropy, whereas the slope reflects the effect of mLOY on heart disease adjusted for horizontal pleiotropy ^32^. Next, we used Mendelian Randomization Pleiotropy Residual Sum and Outlier (MR-PRESSO), an approach that effectively accounts for horizonal pleiotropy by identifying IVs that are outliers ^30^. By doing so, the method prevents potential horizonal pleiotropy and corrects the causal estimates ^30^. Lastly, we used Mendelian Randomization using a Robust Adjusted Profile Score (MR-RAPS), which maximizes the profile likelihood of the ratio estimates while accounting for weak instrument bias, pleiotropy and extreme outliers ^33^. We used the MendelianRandomization, MR-PRESSO and mr.raps packages in R statistical software (version 4.2.2) to conduct MR analyses with summary GWAS data from a previously published study and newly conducted analyses ^1, 34^ (*Online Supplementary Data File*).

## Results

### Baseline characteristics

The study population characteristics at baseline are shown in Table 1. Here, 19.4% of the male study participants had detectable mLOY, while 6.1% of the female study participants had detectable mLOX. On average, men with detectable mLOY were older (61.6 vs. 55.1 years), had higher proportions of former (42.6% vs. 35.8%) or current smokers (16.7% vs. 11.5%), had lower proportions of non-Europeans, and had lower proportions of college/university degree holders (30.4% vs. 35.7%) (Table 1). Similarly, among women, those with detectable mLOX were older (60.6 vs. 55.9 years) on average, had higher proportions of former smokers (34.09% vs. 30.90%), had lower proportions of non-Europeans, and had lower proportions of college/university degree holders (25.6% vs. 32.0%) (Table 1).

### Incident heart disease cases

Among men, there were 14,459 incident cases (hospitalizations) of atrial fibrillation, 6,007 cases of myocardial infarction, 19,507 cases of ischemic heart disease, 1,017 cases of cardiomyopathy, and 5,982 cases of heart failure syndrome during the follow-up (Supplementary Table 1). Among women, there were 9,094 incident cases of atrial fibrillation, 2,609 cases of myocardial infarction, 11,322 cases of ischemic heart disease, 565 cases of cardiomyopathy, and 3,896 cases of heart failure syndrome during the follow-up (Supplementary Table 1). The ages at first diagnosis (hospitalization) and the follow-up time to first diagnosis for each heart disease are shown in Supplementary Table 1.

### Associations of age and smoking with mLOY and mLOX

We conducted unadjusted analyses with age, smoking status, and sex chromosome loss at baseline to check the consistency with previous studies. Age was associated with increased odds of mLOY in men (OR=1.13 per year, 95%CI: 1.13-1.13, p<0.0001) and mLOX in women (OR=1.09 per year, 95%CI: 1.09-1.10, p<0.0001). As expected, among men, former-smokers (OR=1.54, 95%CI:1.50-1.58, p<0.0001) and current-smokers (OR=1.88, 95%CI: 1.82-1.95, p<0.0001) had increased odds of mLOY compared to never-smokers. Among post-menopausal women, former-smoking (OR=1.07, 95%CI: 1.03-1.11, p=0.0014) was associated with increased odds of mLOX, but not current-smoking (OR=0.97, 95%CI: 0.90-1.04, p=0.3865), which was similar to previous analyses ^21^.

### mLOY among men and future risk of heart disease

Among men, having detectable mLOY in leukocytes was significantly associated with elevated risk of incident atrial fibrillation requiring hospitalization (HR=1.06, 95%CI:1.03-1.11, p=0.0011; Table 2), even after accounting for multiple comparisons. Further, the association with atrial fibrillation strengthened non-monotonically as the mosaic CF increased above 20% (HR _CF=0.20-<0.30_=1.20, 95%:1.07-1.35; HR _CF=0.30-<0.40_=0.95, 95%:1.12-1.32; HR_CF=0.40-<0.50_=1.38, 95%:1.13-1.69, p=0.0015; HR_CF>0.50_=1.44, 95%:1.15-1.80, p=0.0015; Figure 1). The association between detectable mLOY and atrial fibrillation was consistent across subgroups including those of European genetic ancestry (HR=1.08, 95%CI:1.04-1.12, p=0.0002), >60 years of age (HR=1.08, 95%CI:1.04-1.13, p=0.0003), and ≤60 years of age (HR=1.07, 95%CI:1.02-1.11, p=0.0021; Supplementary Table 2). The atrial fibrillation associations among never-smokers (HR=1.07, 95%CI:1.01-1.14, p=0.0294) and ever-smokers (HR=1.05, 95%CI:1.01-1.11, p=0.0298) were apparent, but non-significant after Bonferroni-correction. (Supplementary Table 2). Consistent associations were not found for myocardial infarction, ischemic heart disease / angina pectoris, cardiomyopathy, and heart failure syndrome in the overall (Table 2) and stratified analyses (Supplementary Table 2)

**Figure 1:**
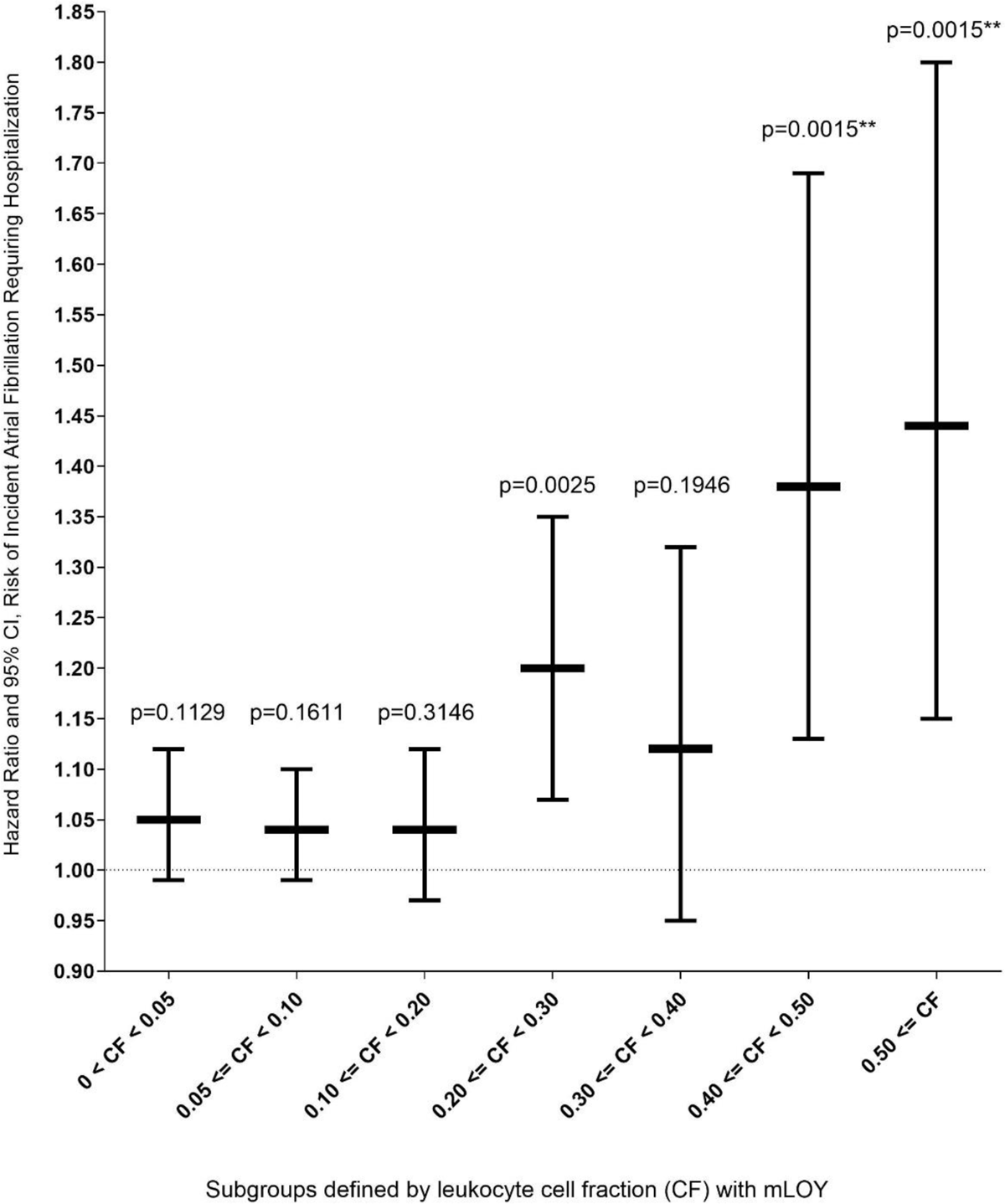
Mosaic loss of chromosome Y (mLOY) in pre-diagnostic leukocytes and risk of newly diagnosed atrial fibrillation, stratified by cell fractions (CF) with mLOY. We analyzed participants with complete genotyping and covariate data. Multivariable Cox regression models were used to estimate hazard ratios (HR) and 95% confidence intervals (CI) of newly diagnosed (incident) atrial fibrillation in relation to mLOY (detectable vs. not detectable) among men. We adjusted for potential confounders including study assessment center, age at recruitment (continuous), self-identified race/ethnicity (European, Black/African, mixed, South Asian, and other groups), 10 principal components for genetic ancestry, educational attainment (i.e., qualifications), smoking status (never, former, current), body mass index (BMI; <18.5, ≥18.5 to <25, ≥25 to <30, ≥30 to <35, and ≥35 kg/m^2^), material deprivation (Townsend deprivation index, socioeconomic status, continuous), alcohol intake (never, former, current occasional, current <1 drink/day, current 1-3 drinks/day, current >3 drinks/day, unknown), diabetes status (none, diabetic, unknown), glycated hemoglobin (HbA1c, mmol/mol, continuous), and hypertension status based on average systolic and diastolic blood pressure at baseline (normal, elevated, stage 1 and 2 hypertension, hypertensive crisis, and unknown), total white blood cell count at enrollment, and any cancer diagnosis at enrollment. Analytic strata sizes with complete data: No detectable mLOY (comparison group): 10,245 cases/153,367 men; 0 <CF<0.05: 1090 cases/11,531 men; 0.05 ≤CF<0.10: 1649 cases/14,905 men; 0.10 ≤CF<0.20: 843 cases/6703 men; 0.20 ≤CF<0.30: 299 cases/1972 men; 0.30 ≤CF<0.40: 141 cases/939 men; 0.40 ≤CF<0.50: 98 cases/533 men; CF<0.50: 78 cases/418 men.

**Table 2:**
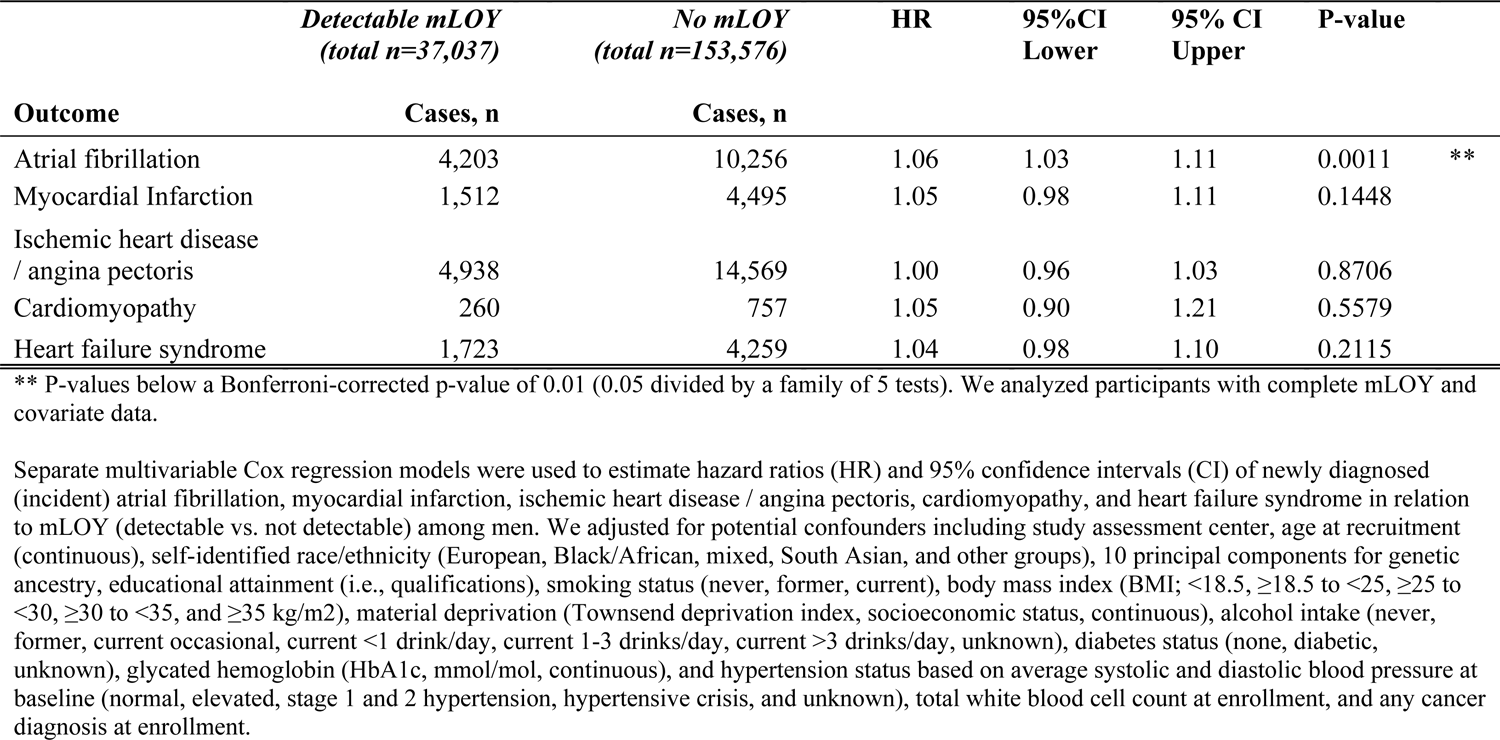
Mosaic loss of chromosome Y (mLOY) in pre-diagnostic leukocytes and risk of incident heart diseases in the UK Biobank.

### Sensitivity and attenuation analyses

To tightly control potential residual confounding from smoking, we adjusted for a 27-category variable for smoking history and intensity ^12, 15^. Here, the mLOY-atrial fibrillation estimates were not considerably changed (HR=1.06, 95%CI:1.02-1.10, p=0.0016). Additionally, we included a quadratic term for age; however, this parameter was non-significant (p=0.19) and the main mLOY-atrial fibrillation association remained statistically significant (HR=1.06, 95%CI:1.02-1.10, p=0.0014).

When analyzing a subgroup of 87,041 participants who had primary care data and met our inclusion criteria, we found that the association between mLOY and general practitioner diagnosed incident atrial fibrillation had a similar effect size but was non-significant (605 cases/16,749 with mLOY; 1470 cases/70,292 without mLOY; HR=1.08, 95%CI: 0.96-1.20, p=0.19) compared to incident hospitalization for atrial fibrillation (1849 cases/16,749 with mLOY; 4538 cases/70,292 without mLOY; HR=1.08, 95%CI: 1.02-1.14, p=8.1×10^-3^). However, there was no evidence that these estimates were different (p-difference _Z score sign_=1.00). When combining general practitioner and hospitalization outcome ascertainment, the association between mLOY and atrial fibrillation risk was statistically significant (1933 cases/16,749 with mLOY; 4746 cases/70,292 without mLOY; HR=1.09, 95%CI: 1.03-1.16, p=2.1×10^-3^).

### Mendelian Randomization, mLOY and atrial fibrillation risk

We conducted MR analyses to assess potential causal associations between mLOY and atrial fibrillation risk among men. Using IVW, we found evidence supporting a causal relationship between mLOY and atrial fibrillation (HR_IVW_=1.15, 95% CI:1.12-1.19, p=1.52×10^-21^; Figure 2). We conducted sensitivity analyses with other MR methods and found that the weighted median (HR_WM_=1.09, 95% CI:1.05-1.14, p=1.42×10^-24^; Figure 2), MR-PRESSO (HR_MR-PRESSO_=1.15, 95% CI:1.13-1.18, p=5.22×10^-5^; Figure 2) and MR-RAPS approaches (HR_MR_=1.10, 95% CI:1.07-1.13, p=1.10×10^-9^; Figure 2) agreed with our main finding. Conversely, the MR-Egger slope was non-significant (p=0.188) and the intercept was non-zero (β_0_= 0.011, p<0.001), suggesting detection of pleiotropy.

**Figure 2:**
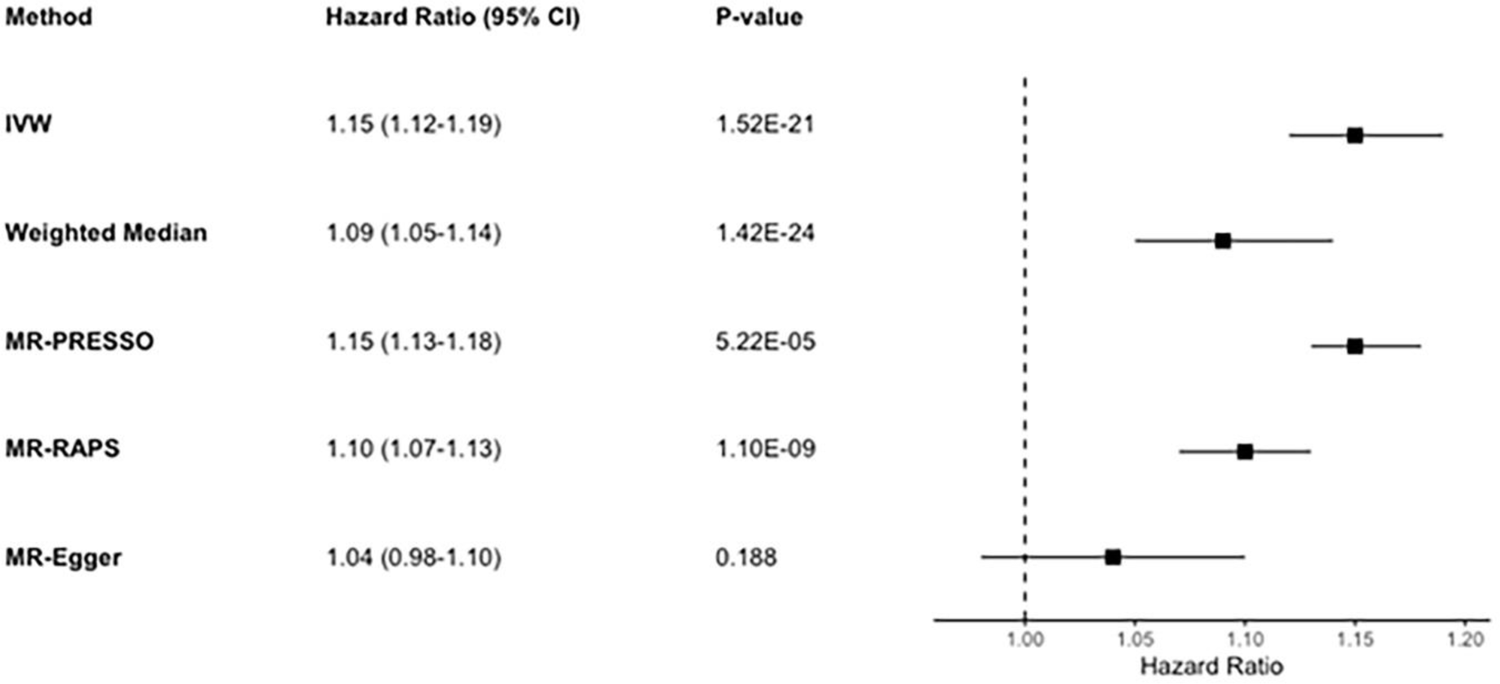
Mendelian Randomization analyses to assess the potential causal associations between pre-diagnostic leukocyte mLOY and risk of newly diagnosed atrial fibrillation in the UK Biobank. CI, confidence interval; IVW, Inverse Variance Weighted; MR-PRESSO, Mendelian Randomization Pleiotropy Residual Sum and Outlier; MR-RAPS, Mendelian Randomization-Robust Adjusted Profile Score.

### mLOX among women and future risk of heart disease

Among women overall, having detectable mLOX in leukocytes was associated with decreased risk of atrial fibrillation requiring hospitalization before accounting for multiple testing; however, this association was marginally non-significant after Bonferroni-correction (HR=0.91, 95%CI:0.84-0.98, p=0.0130; Table 3). The suggestive inverse association between mLOX and atrial fibrillation risk was observed only among post-menopausal women (HR=0.90, 95%CI:0.83-0.98, p=0.0151; Table 3), who comprise most of the female participants. When further stratifying women into subgroups, the suggestive inverse association with atrial fibrillation was consistent among participants of European ancestry (HR=0.91, 95%CI:0.84-0.98, p=0.0145), and those above and below 60 years of age at baseline (HR=0.90, 95%CI:0.83-0.98, p=0.0164 and HR=0.89, 95%CI:0.82-0.96, p=0.0044, respectively) before accounting for multiple testing, but was significant only among women ≤60 years of age after Bonferroni-correction (Supplementary Table 3).

**Table 3:**
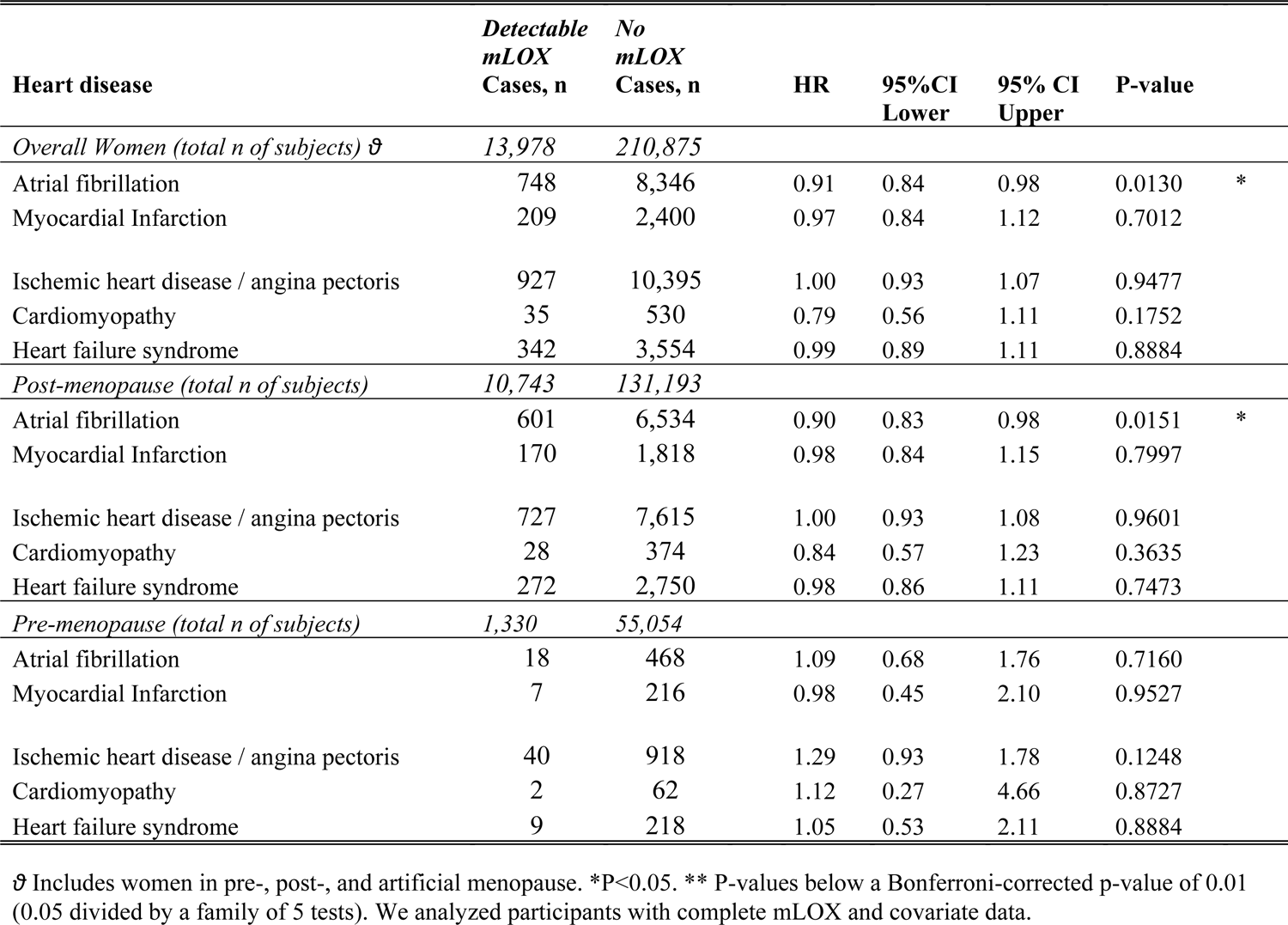
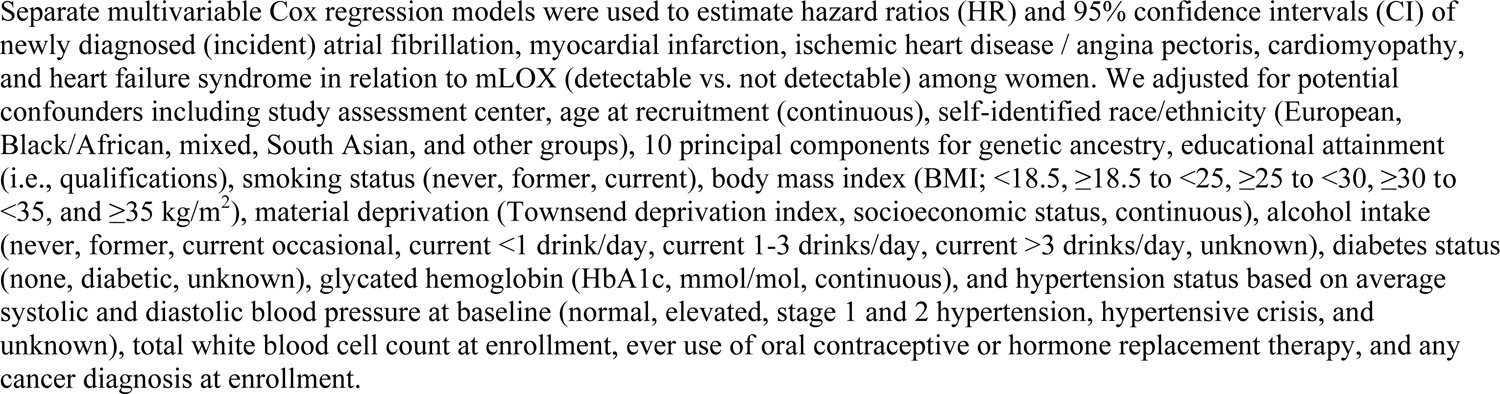
Mosaic loss of chromosome X (mLOX) in pre-diagnostic leukocytes from women and risk of incident heart diseases in the UK Biobank.

## Discussion

In a large prospective cohort study from the United Kingdom, we investigated the associations between mLOY and mLOX in pre-diagnostic leukocytes collected at baseline and risk of incident heart disease requiring hospitalization. Among men, we found that mLOY was significantly associated with elevated risk of incident atrial fibrillation. This association was greatly strengthened among participants with increased CF with mLOY and was largely consistent across subgroups defined by smoking, age, and race/ethnicity. Further, most of the MR analyses supported causal associations between mLOY and atrial fibrillation. However, consistent associations between mLOY and other heart diseases were not detected. To our knowledge, we conducted the first study of mLOX among women and risk of newly diagnosed heart disease. Here, we found suggestive evidence for inverse associations between mLOX and atrial fibrillation risk that was non-significant after accounting for multiple comparisons. When stratifying by menopausal status, the suggestive association between mLOX and atrial fibrillation was only observed among post-menopausal women. Furthermore, given that mLOX is less frequent among younger women ^19^ and that most female UK Biobank participants were >50 years of age, we did not have sufficient sample size to make sound inferences for pre-menopausal women. Additionally, consistent associations were not detected between mLOX other heart diseases among women.

Our study examines the relationships between sex chromosome mosaicisms and CVD by focusing specifically on incident heart disease hospitalization, as opposed to prevalence and mortality as in other studies ^15, 16, 35^. We provide population-based evidence that mLOY and mLOX could be involved (either directly or indirectly as a surrogate marker) in the biological processes leading up to atrial fibrillation occurrence. Our investigation builds upon previous studies conducted in human populations. In the UK Biobank, Loftfield et al. (2018) reported cross-sectional associations between prevalent heart disease and stroke and increased leukocyte mLOY at baseline ^15^. Further, they reported no evidence for associations between mLOY and mortality from cardiovascular disease, ischemic heart disease, and stroke ^15^. Additionally, a recent study by Sano et al. (2022) found evidence for associations between mLOY and death caused by diseases of the circulatory system including hypertensive heart disease, atrial fibrillation, as well as heart failure syndrome ^16^.

We note that our outcome ascertainment using national hospital registries potentially enriches for more serious cases of heart disease, which can be non-fatal or fatal. As such, we conducted analyses using a subset of participants with preliminary primary care data, in which the cases had moderate, non-fatal incident heart disease. Here, we found that the associations between mLOY and general practitioner diagnosed atrial fibrillation were similar in magnitude to the hospitalization data, but was not statistically significant. This was likely due to limited statistical power from reduced case numbers. Important for consideration, these preliminary primary care data have not been validated or adjudicated and we include these only for exploratory purposes and sensitivity analyses. When combining general practitioner diagnosed and hospitalization cases, the mLOY-atrial fibrillation association was statistically significant.

Our MR analyses using IVW, weighted median, MR-PRESSO, and MR-RAPS strongly supported causal associations between mLOY and atrial fibrillation and had similar causal effect estimates. MR-PRESSO is a newer method that effectively accounts for outlying genetic instruments that show evidence of horizontal pleiotropy; however, no outliers were detected in these analyses. Similarly, the newest method, MR-RAPS, accounts for weak instrument bias, pleiotropy and extreme outliers. However, we found that dose-response relationship between the genetic associations with mLOY and atrial fibrillation was non-significant using the older MR-Egger regression, and the non-zero intercept suggested some pleiotropy. We note that MR-Egger has been reported to be less statistically efficient and subject to biased or skewed estimates ^36^, particularly when the InSIDE (INstrument Strength Independent of Direct Effect; independence between the Egger regression intercept and slope) assumption is violated ^31^.

The precise biological roles of mLOY and mLOX in the pathogenesis of heart disease in humans is unclear, but mechanistic studies using mouse models provide some clues. For example, Sano et al. (2022) demonstrated that mLOY promotes fibrotic phenotypes (‘scarring’) in cardiac murine tissues which potentially impairs cardiac function such as tissue elasticity and electrical signal propagation ^16^. Further, Horitani et al. (2024) found evidence that mLOY in mice may disrupt the ubiquitously transcribed tetratricopeptide repeat containing, Y-linked (Uty) gene, leading to epigenetic dysfunction and differentiation of profibrotic macrophages ^37^. Indeed, this mechanistic evidence from mice is consistent with the association we found between mLOY and atrial fibrillation risk and is consistent with the pathology of atrial fibrillation. Furthermore, evidence from sequencing data suggests that mLOY is genetically linked to clonal hematopoiesis of indeterminate potential (CHIP) ^38–40^, a mosaic expansion of hematopoietic stem cells driven by certain somatic mutations ^41^, which has been recently associated with risk of CVDs and heart failure syndrome ^42–46^. Additionally, it has been hypothesized that mLOY in leukocytes potentially reflects altered immunosurveillance and inflammatory processes, which can influence cardiovascular disease pathogenesis ^3,47^.

We note that atrial fibrillation risk was positively associated with mLOY among men as expected but was potentially inversely associated with mLOX among women. The underlying reason for this potential inverse association with mLOX is unknown. It is possible that mLOY and mLOX operate in a distinctly sex-specific manner in reflecting genomic instability from the cumulative burden of age, genetic susceptibility, and environmental exposures.

Our study had numerous strengths. First, we used MoChA to call chromosomal mosaicisms from GWAS data, which is more sensitive compared to previous calling approaches ^17^. Second, the prospective cohort design strengthened our inferences and is robust against disease-effect bias of pre-diagnostic leukocyte mLOY and mLOX (i.e., “reverse causation”). Third, we analyzed a large sample size of middle-aged and older adults which improved statistical power to detect modest associations.

Our study had some limitations. First, even though our female sample size was large, we had limited power to detect modest associations, particularly among younger pre-menopausal women. However, we did detect suggestive associations for mLOX among post-menopausal women which could serve as a foundation for future analyses in larger pooled studies. Second, although we thoroughly adjusted for measured confounders related to chromosomal mosaicisms and heart disease, we cannot discount the possibility of unmeasured or residual confounding.

However, our findings were consistent across never- and ever-smokers and age groups; supported by MR analyses; robust to adjustment for detailed smoking history; and consistent with previous studies ^16^. Lastly, there is the possibility that healthy volunteer bias in the UK Biobank could limit generalizability; however, this does not affect the internal validity of our mechanistic study.

In conclusion, mLOY was significantly associated with elevated risk of atrial fibrillation among men. Conversely, we found suggestive evidence of an inverse association between mLOX and atrial fibrillation risk among post-menopausal women. Associations were not detected for other heart diseases. Caution is recommended when interpreting the findings, particularly for mLOX among women. Although the precise biological role of sex chromosome mosaicisms in heart disease pathogenesis remains unclear, our findings support biologic processes related to or tagged by leukocyte mLOY and mLOX are related to atrial fibrillation occurrence in human populations. If our findings are confirmed across diverse populations, they support further consideration of mLOY in atrial fibrillation risk prediction and stratification analyses among men.

## Data Availability

The UK Biobank data is publicly available at: https://ams.ukbiobank.ac.uk/ams/. Metadata are available are available upon reasonable request.

## Acknowledgements

This study was supported by intramural funding from the National Cancer Institute and National Heart, Lung, and Blood Institute. We thank Drs. Qing Lan and Nathaniel Rothman for their scientific support. We extend our appreciation to Zhonghua Liu, Haoyu Zhang, and Peter Kraft for their advice on interpreting the MR analyses. Additionally, we thank Lisa Finkelstein, Jillian Varonin, and the UK Biobank Access Team for helping us with international data agreements. We declare no conflicts of interest. Lastly, we thank Giulio Genovese and John R.B. Perry for contributing to the calling of chromosomal mosaicisms.

---

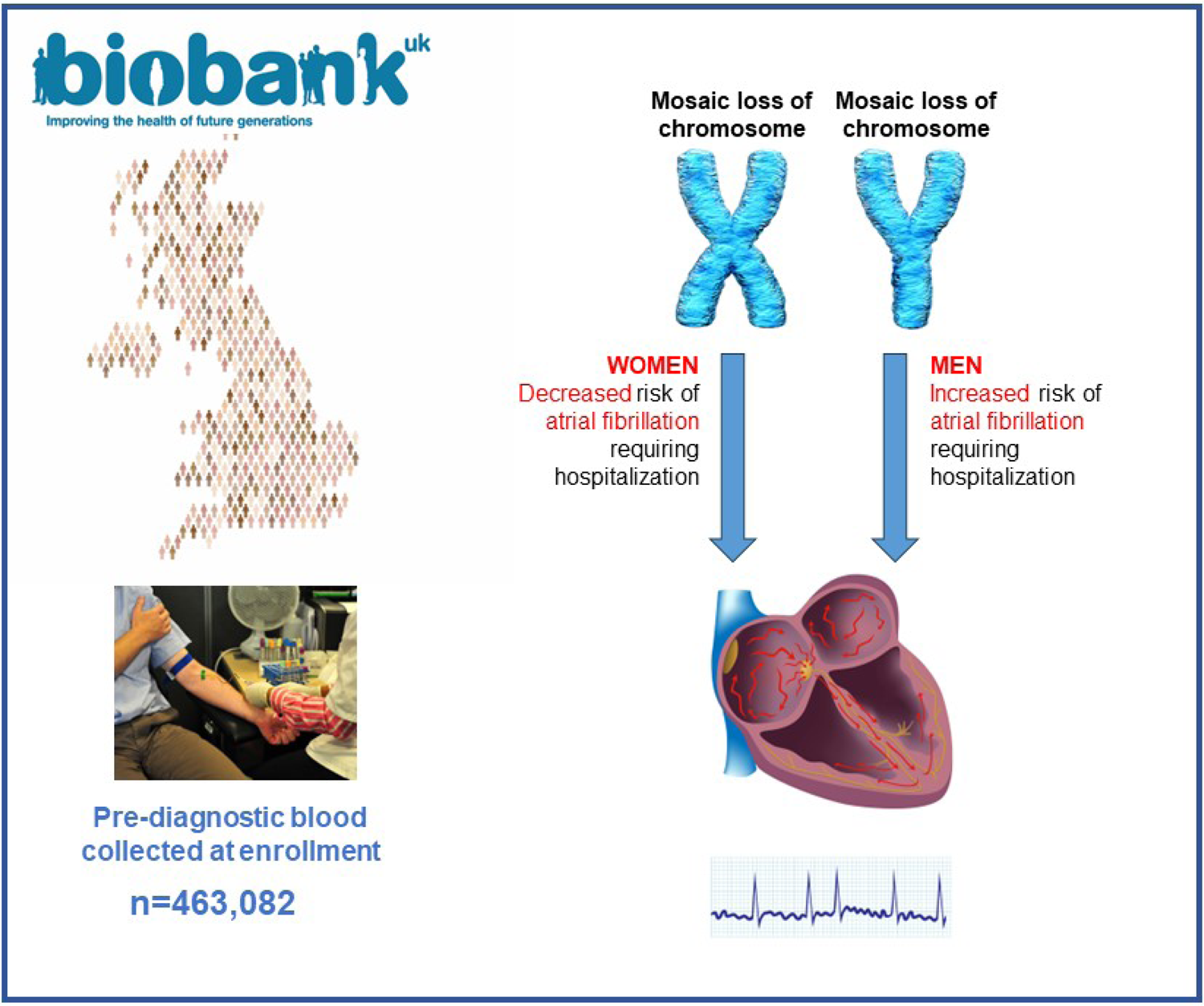

